# Molecular Underpinnings of Retinal Traits Shared with Major Psychiatric Disorders

**DOI:** 10.64898/2026.08.31.26361809

**Authors:** Piotr Jaholkowski, Nadine Parker, Imre Odin Sveen, Erik D Wiström, Vera Fominykh, Attila Szabo, Pravesh Parekh, Oleksandr Frei, Olav B. Smeland, Kevin S O’Connell, Srdjan Djurovic, Anders M. Dale, Alexey A. Shadrin, Ole A. Andreassen

## Abstract

Recent large-scale studies have enabled new knowledge about genetic underpinnings of morphological and electrophysiological alterations of the retina. Variation in retinal traits, often of neurodevelopmental origin, have been linked to major psychiatric disorders (MPDs). Here, we investigate the genetic overlap between MPDs and key retinal traits to identify underlying molecular mechanisms. We obtained genome-wide associations studies data for bipolar disorder (BD), major depression (MD), schizophrenia (SCZ), and the retinal traits retinal nerve fibre layer thickness (RNFL), ganglion cell inner plexiform layer thickness (GCIPL), and vertical cup-disc ratio (VCDR). We estimated the number of trait-influencing variants shared between traits with MiXeR and identified shared genetic loci with condFDR. Subsequently, we examined the biological pathways of the genes mapped to shared loci. This revealed that GCIPL shared the most genetic variants with MPDs (∼60%), followed by RNFL (∼40%), and VCDR (∼20%). The genetic variants shared between retinal traits and MPDs showed disorder-specific patterns with more pronounced overlaps of SCZ and BD with RNFL, and MD negatively correlated with GCIPL. Gene-pathway analysis highlighted the importance of GABAergic neurotransmission and a two-stage neurodevelopmental process in SCZ, whereas the role of mitochondria and a weaker developmental component were observed in BD. The results also implicated synaptic functioning and gene-expression processes in MD. Furthermore, polygenic analysis suggested that the genetic architecture of retinal traits can distinguish between MPDs. Our findings indicate shared genetic underpinnings between retinal traits and SCZ, BD, and MD, implicating altered neurodevelopment and neurotransmission underlying the retinal link to major psychiatric disorders.

## Introduction

Histological studies have revealed differences in the number and structure of brain cells in major psychiatric disorders (MPDs) such as schizophrenia (SCZ)(1, 2), bipolar disorder (BD)(3), and major depression (MD)(4). Notably, these changes exhibit regional, cellular, and neurotransmitter-type specificity and seem to have disorder-specific patterns(1–4). Thus, the evaluation of structural brain alterations may elucidate common and disorder-specific pathophysiological mechanisms across MPDs and improve diagnostic differentiation. However, histological studies of MPDs are constrained by small sample size, limited representativeness of specimens due to donors with more severe clinical manifestations and advanced age, and the fundamental lack of in vivo assessments(1, 3).

The retina, which can be observed noninvasively, provides a unique window into the structure and function of the central nervous system (CNS). The retina displays a highly ordered, multilaminar organization composed of specialized neuronal networks(5). Also its physiology, encompassing a broad repertoire of neurotransmitters, an immune-privileged microenvironment, and responses to insult, mirrors that of other CNS regions(5). The most commonly studied retinal traits are thickness of the ganglion cell axon layer (retinal nerve fiber layer, RNFL)(6), the thickness of the ganglion cell body layer (ganglion cell inner plexiform layer, GCIPL)(6), and a quantitative indicator for optic nerve head morphology (vertical cup-disc ratio, VCDR)(7).

In MPDs, a series of retina studies have indicated morphological, biochemical, and electrophysiological alterations(5, 8–17). It has been demonstrated that SCZ, BD, and MD are associated with structural changes in the retinal layers(8, 18–20) and that these disorders differ in the pattern of changes in specific retinal layers and regions(8). Electroretinography studies have revealed differences in retinal function between SCZ, BD, or MD and healthy controls(21, 22), as well as differences between SCZ and BD that may help improve discrimination between these disorders(21). Moreover, the neurodevelopmental nature of these electroretinographic anomalies is indicated by their occurrence in healthy children and adolescents at genetic risk for psychosis(23). Additionally, reduced cone system amplitudes, predominantly in the central retina, together with reduced oscillatory potentials in SCZ, indicate disturbed dopaminergic retinal transmission(24). The genetic factors underlying these retinal changes in MPDs remain elusive.

The evaluation of the genetic architecture shared between MPDs and retinal traits has recently become feasible due to large-scale genome-wide association studies, which indicate that, akin to MPDs(25–27), retinal traits such as the RNFL(6), GCIPL(6), and VCDR(7) are also under polygenic influences. Here we investigate whether retinal traits exhibit genetic overlap with MPDs, to determine if the associations from clinical studies have a genetic basis. Further, we investigate whether the shared genetic architectures with retinal traits are specific to each disorder (SCZ, BD, MD). Previous research has demonstrated that the genetic architecture shared among human traits often comprises a balanced mixture of genetic variants with both concordant and discordant effect directions(28, 29), which may underlie the weak genetic correlations previously observed(30). To address these challenges in studying genetic architecture, we applied the bivariate causal mixture model (MiXeR), which estimates the number of trait-influencing genetic variants shared between two traits and the genetic correlation of these shared variants, irrespective of the genetic correlation between the traits(28, 31). Similarly, the conjunctional false discovery rate (conjFDR) approach can identify genetic loci shared between traits and disorders irrespective of effect direction(32). We then performed extensive follow-up analysis to obtain more insight into the neurobiological mechanisms shared between the retina and MPDs.

## Methods

### GWAS summary statistics

The GWAS summary statistics for the retinal nerve fibre layer thickness (RNFL; 31,434 individuals)(6), the ganglion cell inner plexiform layer thickness (GCIPL; 31,434 individuals)(6), and vertical cup-disc ratio (VCDR; 65,680 individuals)(7) were obtained from the NHGRI-EBI GWAS Catalog(33). The GWAS summary statistics for schizophrenia (SCZ; 53,386 cases and 77,258 controls)(27), bipolar disorder (BD; 59,287 cases and 781,022 controls)(26), and major depression (MD; 412,305 cases and 1,588,397 controls)(25) were obtained from the Psychiatric Genomics Consortium(34). To avoid sample overlap in the conjFDR analyses, we also acquired summary statistics for BD and MD after excluding the UK Biobank sample. The GWASs participants were predominantly of European ancestry. All GWAS investigated in the present study were approved by the relevant ethics committees, and informed consent was obtained from all participants. The Regional Committee for Medical and Health Research Ethics for the South-East Norway region evaluated the protocol concerning the use of GWAS summary statistics and concluded that no additional institutional review board approval was required, as no individual-level data were used.

### TOP Study

The naturalistic Thematically Organized Psychosis (TOP) Study includes participants aged 18-65 years recruited from psychiatric units of Oslo hospitals, along with healthy controls form the same catchment area(35). All participants provided written informed consent, and the study was approved by The Regional Committee for Medical and Health Research Ethics of South-East Norway.

In the current study we included a subsample of the TOP cohort comprising individuals with diagnosis of SCZ (n=707), BD (n=920) or MD (n=260) according to the Diagnostic and Statistical Manual of Mental Disorders, Fourth Edition (DSM-IV)(36). The analyzed group was restricted to the individuals of European ancestry. For each related pair of study individuals with a kinship coefficient greater than 0.05 (PLINK2 v00a6LM (37)), we excluded one member, prioritizing the retention of MD cases, secondarily, SCZ cases.

### Modeling trait-influencing variants using MiXeR

We applied the statistical tool MiXeR v1.3 to the GWAS summary statistics to evaluate the polygenic overlap across the phenotypes included in our study(31, 38). Univariate causal mixture models were used to estimate the number of trait-influencing variants (polygenicity_90_) explaining 90% of the SNP heritability (*h²_SNP_*) for each phenotype(38). We utilized bivariate causal mixture models (bivariate MiXeR) for pairs of phenotypes to estimate the number of trait-influencing variants shared between the phenotypes, unique to phenotype 1, and unique to phenotype 2, irrespective of the genetic correlation between them(31). Moreover, we derived the genome-wide genetic correlation (*r_g_*) and the genetic correlations of the variants shared between phenotypes (*r_gs_*) from the bivariate MiXeR models.

### Estimating shared variants with conjFDR

To visualize cross-trait SNP enrichment among the pairs of retinal traits and MPDs, we constructed QQ-plots illustrating the distribution of *p*-values for the primary trait conditioned on significance levels in the secondary trait and vice versa(32). We controlled for spurious enrichment by generating the QQ-plots after random pruning, averaging over 500 iterations(32). For each iteration, we removed all but one random SNP in each linkage disequilibrium (LD) independent region (clump of SNPs in strong LD, *r^2^*>0.1). Moreover, to prevent bias, SNPs within long-range linkage disequilibrium (LD) regions that show strong associations with brain traits. Successive leftward deflections of a primary phenotype on a QQ-plot, associated with increasing levels of association with a secondary phenotype, were used as an indicator of cross-trait enrichment(32).

We used the conjFDR approach to identify genetic loci shared between retinal traits and MPDs(32). The conjFDR approach is based on running two conditional (condFDR) analysis that recalculate the associations between variants and a primary phenotype based on their associations with the secondary phenotype and vice versa. Subsequently, the higher of the two obtained condFDR values for a specific variant is taken as the conjFDR, which provides a conservative estimate of the association between that variant and the two phenotypes of interest(32). We applied an FDR significance cutoff of 0.05 for our conjFDR analyses. The cond/conjFDR analyses were performed after removing genomic regions with a complex LD pattern (chr6:25119106-33854733; chr8:7200000-12500000; chr19:44909039-45912650).

Independent genomic loci were identified according to the FUMA protocol(39). We defined independently significant SNPs as those with conjFDR<0.05 and an LD *r²*<0.6 with each other. Subsequently, we identified independent significant lead SNPs as those with LD *r²*<0.1 with each other. The applied boundaries of each genomic locus included SNPs with an LD *r²*≥0.6 relative to any of the independently significant SNPs within the locus. We merged loci separated by less than 250 kb. The lead SNP was defined as one having the lowest conjFDR within a given locus. The 1000 Genomes Project European ancestry haplotype reference panel was used to compute LD *r²*-values(40).

### Functional annotation

To map genes to lead SNPs identified in the conjFDR analyses, we used positional information provided by the Open Targets Platform (access:17/07/2025)(41). Subsequently, for each MPD separately, we combined the lists of genes shared between a respective MPD and the three retinal traits. Then, using BrainSpan RNA sequencing data and the *cerebroViz* R package, we plotted heatmaps visualizing the spatiotemporal expression of the genes(42, 43). Moreover, we used nonlinear LOESS modeling implemented in the *ggplot2* R package to visualize mean expression across lifespan periods in cortical structures, subcortical structures, and both groups combined(44).

Similarly, we merged the list of genes associated with specific MPD traits and the three retinal traits, and we utilized the Whole Human Brain 10x RNA-seq gene expression data to visualize gene expression in cells belonging to different taxonomic groups(45). We followed the data preparation and clustering procedures outlined by the Allen Institute and visualized the heatmaps of gene expression for specific cell clusters using the *pheatmap* R package for each MPD(45–47).

The gene set enrichment analyses were conducted for a combined set of genes mapped to all independently significant SNPs shared between a given MPD and the three retinal traits. For this purpose, we used Open Targets Genetics (access:02/06/2025) to map genes for independent significant SNPs (including lead SNPs) from conjFDR analyses using positional information and an overall score, which is an aggregated measure based on positional information, chromatin interactions, quantitative trait loci, and in silico functional prediction datasets(48). Specifically, for each independent significant SNP, we selected the nearest gene, along with any genes that had an overall score equal to or higher than that of the nearest gene. We run analysis after excluding loci within MHC region. We applied the list of mapped genes to the *g:GOSt* function of the g:Profiler (access:03/06/2025) to identify gene set enrichment within the Gene Ontology (GO) and Kyoto Encyclopedia of Genes and Genomes (KEGG) gene sets(49). We applied the *g:SCZ* method as a multiple testing correction, which is a standard and specifically designed approach for g:Profiler(49).

Moreover, to assess interactions between genes mapped to loci shared between retinal traits and MPDs, we used PCNet v2.2(50), which integrates information on interactions such as protein-protein interactions, gene regulation, signaling, and colocalization. The protein-coding status of genes was evaluated using the GENECODE database v49lift37(51). To avoid bias related to interaction between gene located next to each other on the DNA strand, for each locus we included only the gene nearest to the lead SNP. For same reason, we merged all overlapping loci identified for a given MPD and retinal traits and used, in the analysis, the lead SNP with the lowest conjFDR within a merged locus. Subsequently, we assess the number of interactions between mapped genes, as well as the number of genes interacting with each other within a given gene set. To evaluate statistical enrichment of those two parameters, we compared them with those obtained for 50,000 randomly generated gene sets of the same size, drawn from the GENCODE protein-coding gene set(51). We reported one-sided nominal *p*-values (*p*_nominal_) and *p*-values corrected for multiple testing using the Benjamini-Hochberg procedure (*p*_adjusted_) for each parameter separately. We visualized gene product networks using the Cytoscape v3.10.4(52) and *py4cytoscape* v1.13.0(53) Python package.

### Polygenic risk score analysis

To investigate the relationship between the genetic underpinnings of retinal traits and MPD diagnoses in a case-case comparison, we conducted PRS analyses in an independent cohort of individuals with MPD diagnoses (TOP).

We applied PRSice v2.3.5(54) to calculate the PRSs from GWAS of RNFL(6), GCIPL(6), and VCDR(7). Subsequently, following a widely applied approach (55–57), we extracted the first principal component for each PRS across *p*-value thresholds (5.0e-08, 1.0e-06, 1.0e-05, 1.0e-04, 1.0e-179 03, 1.0e-02, 5.0e-02, 1.0e-01, 5.0e-01, 1.0).

Logistic regression was used to test the association between each of the three retinal PRSs and diagnostic group (i.e. SCZ, BD, and MD) in case-case comparisons, estimating the association between each PRS and one diagnostic category versus another. Covariates included the first 10 genome-wide principal components, sex and age. We reported the nominal *p*-value (*p*_nominal_) and the *p*-value corrected for multiple testing using the Benjamini-Hochberg correction (*p*_adjusted_).

## Results

### Modeling trait-influencing variants using MiXeR

Consistent with prior studies, the most polygenic trait among the assessed psychiatric disorders was MD (polygenicity_90_=11770 trait-influencing variants explaining 90% of MD’s *h^2^_SNP_*, SD=360), followed by SCZ (polygenicity_90_=9540 trait-influencing variants explaining 90% of SCZ’s *h^2^SNP*, SD=220) and BD (polygenicity =7780 trait-influencing variants explaining 90% of BIP’s *h^2^SNP*, SD=240). In contrast, retinal traits demonstrated substantially lower polygenicity, with the most polygenic trait being VCDR (polygenicity_90_=820 trait-influencing variants explaining 90% of VCDR’s *h^2^_SNP_*, SD=31), followed by RNFL (polygenicity_90_=570 trait-influencing variants explaining 90% of RNFL’s *h^2^_SNP_*, SD=64) and GCIPL (polygenicity_90_=470 trait-influencing variants explaining 90% of GCIPL’s *h^2^_SNP_*, SD=68) (Supplementary Table 1).

Using bivariate MiXeR analysis, we estimated that for CGIPL, 55%, 70%, and 70% of trait-influencing variants were shared with SCZ, BD, and MD, respectively (Figure 1, Supplementary Table 1). In the case of RNFL, 40%, 45%, and 40% of trait-influencing variants were shared with SCZ, BD, and MD, respectively. For VCDR, 15%, 30%, and 20% of trait-influencing variants were shared with SCZ, BD, and MD, respectively. We estimated that GCIPL shared 88% of trait-influencing variants with RNFL, while VCDR shared 19% and 38% of trait-influencing variants with GCIPL and RNFL, respectively.

**Figure 1.**
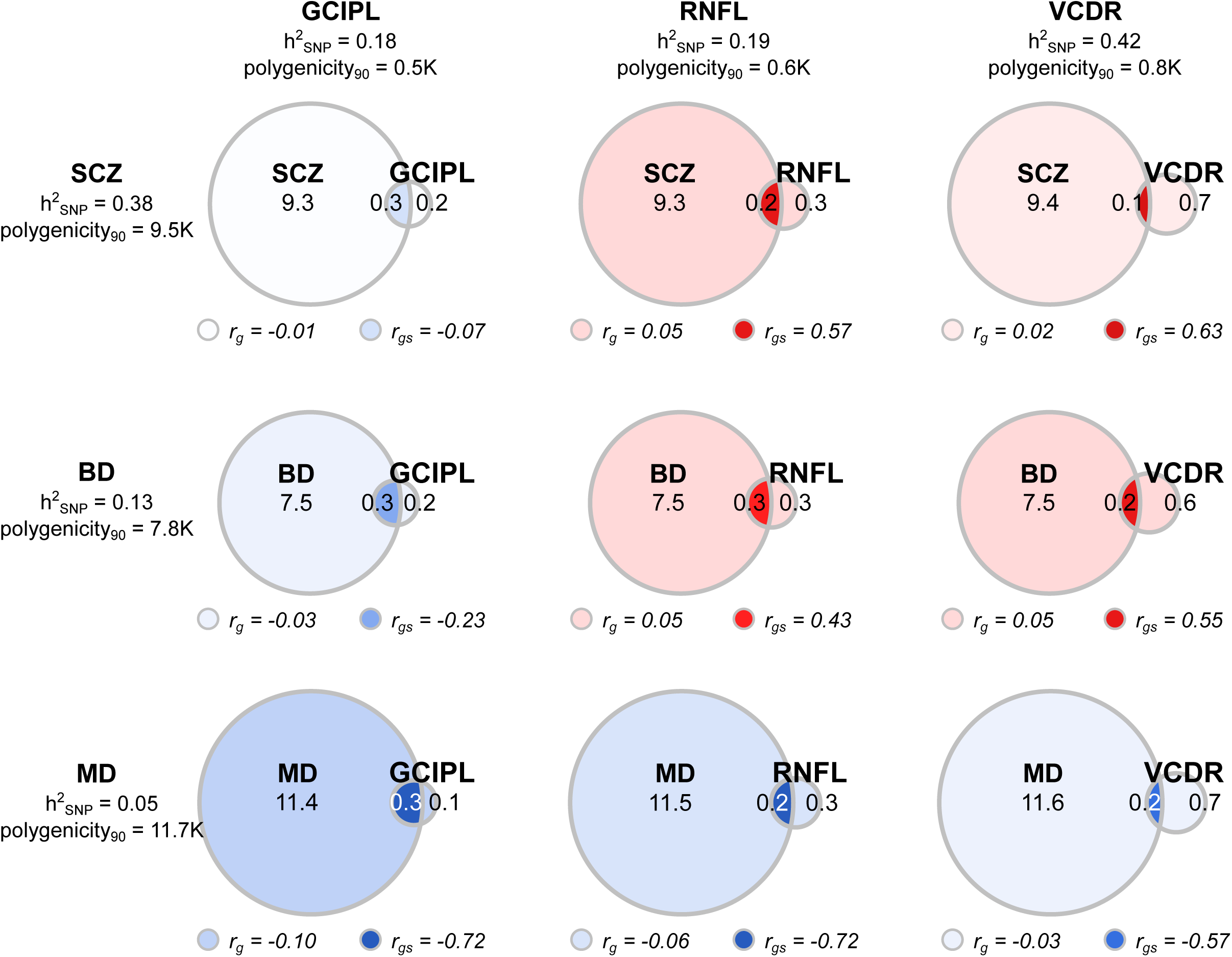
Venn diagrams illustrating MiXeR-modeled genetic overlap, genome-wide genetic correlation, and genetic correlation of shared variants between retinal traits and major psychiatric disorders. The size of the circles reflects the polygenicity of each trait. The unique and shared regions of the Venn diagrams show the numbers of unique and shared trait-influencing variants, respectively, with the number of trait-influencing variants shown in thousands. Genome-wide genetic correlations (r_g_) are depicted below each Venn diagram and are represented by coloring the unique parts of each diagram. Genetic correlations of shared variants (r_gs_) are depicted below each Venn diagram and are represented by coloring the shared part of each diagram. The colours illustrate correlations, ranging from 1 (dark red) to −1 (dark blue). Additionally, the SNP-based heritability (h²_SNP_) and total polygenicity (polygenicity_90_) are presented for each trait. Ganglion cell inner plexiform layer thickness (GCIPL); retinal nerve fibre layer thickness (RNFL); vertical cup-disc ratio (VCDR); schizophrenia (SCZ); bipolar disorder (BD); major depression (MD).

GCIPL showed a weak negative correlation with all disorders - SCZ, BD, and MD - at the genome-wide level (*r_g_*=−0.01 [*SD*=0.01], *r_g_*=−0.03 [SD=0.02], *r_g_*=−0.1 [*SD*=0.01], respectively) (Figure 1, Supplementary Table 1). While the shared variants between GCIPL and MD were strongly negatively correlated (*r_gs_*=−0.72 [*SD*=0.19], those between GCIPL and BD, as well as between GCIPL and SCZ, were weakly negatively correlated (*r_gs_*=−0.23 [*SD*=0.23] and *r_gs_*=−0.06 [*SD*=0.14], respectively).

At the genome-wide level, RNFL showed a weak correlation with SCZ and BD (*r_g_*=0.05 [*SD*=0.01] and *r_g_*=0.05 [*SD*=0.02], respectively), and a weak negative correlation with MD (*r_g_*=−0.06 [*SD*=0.01]) (Figure 1, Supplementary Table 1). The shared variants between RNFL and SCZ, as well as between RNFL and BD, were moderately correlated (*r_gs_*=0.57 [SD=0.22] and *r_gs_*=0.43 [*SD*=0.17], respectively), while those between RNFL and MD were strongly negatively correlated (*r_gs_*=−0.72 [*SD*=0.24]).

VCDR, at the genome-wide level, showed a weak correlation with SCZ and BD (*r_g_*=0.02 [*SD*=0.01] and *r_g_*=0.05 [*SD*=0.01], respectively), and a weak negative correlation with MD (*r_g_*=−0.03 [*SD*=0.01]) (Figure 1, Supplementary Table 1). The shared variants between VCDR and SCZ, as well as between VCDR and BD, were moderately correlated (*r_gs_*=0.63 [*SD*=0.21] and *r_gs_*=0.55 [*SD*=0.14], respectively), while those between VCDR and MD were moderately negatively correlated (*r_gs_*=−0.57 [*SD*=0.23]).

GCIPL showed moderate genetic correlation with RNFL at the genome-wide level as well as at the level of the shared variants (*r_g_*=0.46 [*SD*=0.02] and *r_gs_*=0.65 [*SD*=0.09]) (Figure 1, Supplementary Table 1). VCDR, at the genome-wide level, showed a weak negative correlation with GCIPL and RNFL (*r_g_*=−0.11 [*SD*=0.01] and *r_g_*=−0.01 [*SD*=0.01], respectively). The shared variants between VCDR and GCIPL were moderately correlated (*r_gs_*=−0.45 [*SD*=0.12]), while those between VCDR and RNFL were weakly negatively correlated (*r_gs_*=−0.03 [*SD*=0.03]).

### Estimating shared variants with conjFDR

ConjFDR analysis at FDR<0.05 identified 12 loci shared between SCZ and GCIPL, 2 loci shared between BIP and GCIPL, as well as 14 loci shared between MD and GCIPL (Figure 2, Supplementary Table 2-3). Concordant effect direction was observed for 4 (33.33%) loci shared between SCZ and GCIPL, for 8 (57.14%) loci shared between MD and GCIPL. Both loci shared between BD and GCIPL had discordant effect direction.

**Figure 2.**
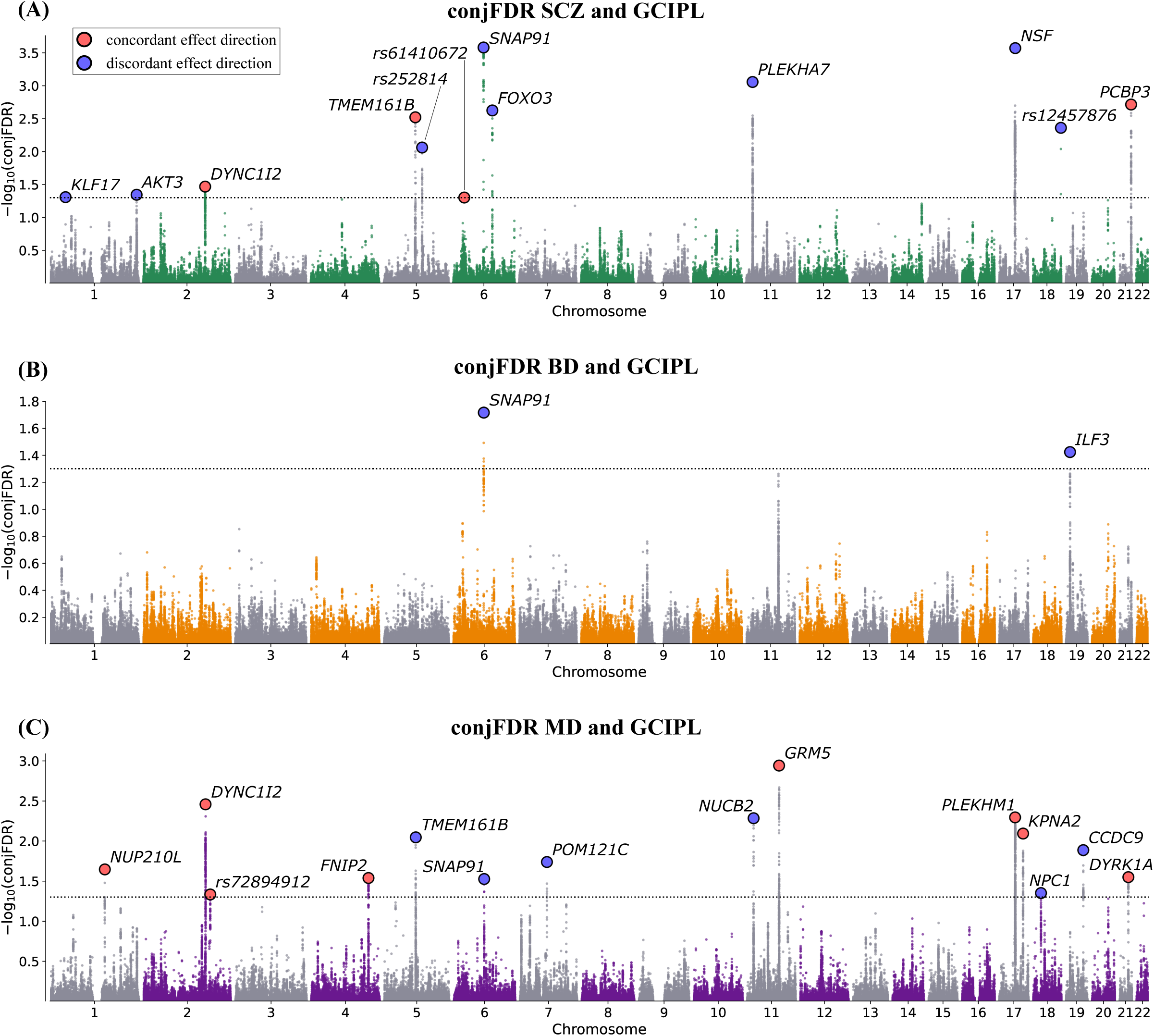
The conjFDR Manhattan plots illustrate common genetic variants that are jointly associated with **(A)** schizophrenia (SCZ) and ganglion cell inner plexiform layer thickness (GCIPL), **(B)** bipolar disorder (BD) and GCIPL, and **(C)** major depression (MD) and GCIPL at conjunctional false discovery rate (conjFDR)<0.05. The y-axis shows the −log10 transformed conjFDR. Chromosomal position is presented along the x-axis. The threshold for significant shared associations (conjFDR<0.05) is represented by the horizontal dotted line. Lead SNPs are indicated by a black outline. Lead SNPs with concordant effect directions for both traits are depicted in red, whereas those with discordant effect directions are depicted in blue.

Using conjFDR analysis (FDR<0.05) we identified 14 loci shared between SCZ and RNFL, 2 loci shared between BIP and RNFL, as well as 9 loci shared between MD and RNFL (Supplementary Figure 5, Supplementary Table 5-7). Concordant effect direction was observed for 6 (42.86%) loci shared between SCZ and RNFL, for 7 (77.78%) loci shared between MD and RNFL. Both loci shared between BD and RNFL had concordant effect direction.

ConjFDR analysis at FDR<0.05 identified 23 loci shared between SCZ and VCDR, 15 loci shared between BIP and VCDR, as well as 35 loci shared between MD and VCDR (Supplementary Figure 6, Supplementary Table 8-10). Concordant effect direction was observed for 13 (56.52%) loci shared between SCZ and VCDR, for 8 (53.34%) loci shared between BD and VCDR, and for 15 (42.86%) loci shared between MD and VCDR.

Gene expression analysis in the Whole Human Brain Atlas 10X RNA-seq dataset demonstrated that genes shared between retinal traits and all MPDs are predominantly expressed in cells belonging to clusters containing neuronal cells (Figure 3A, C, E). Furthermore, this expression occurs at similar levels in cells belonging to different neuronal cell clusters, indicating a lack of specificity for neuronal cell clusters.

**Figure 3.**
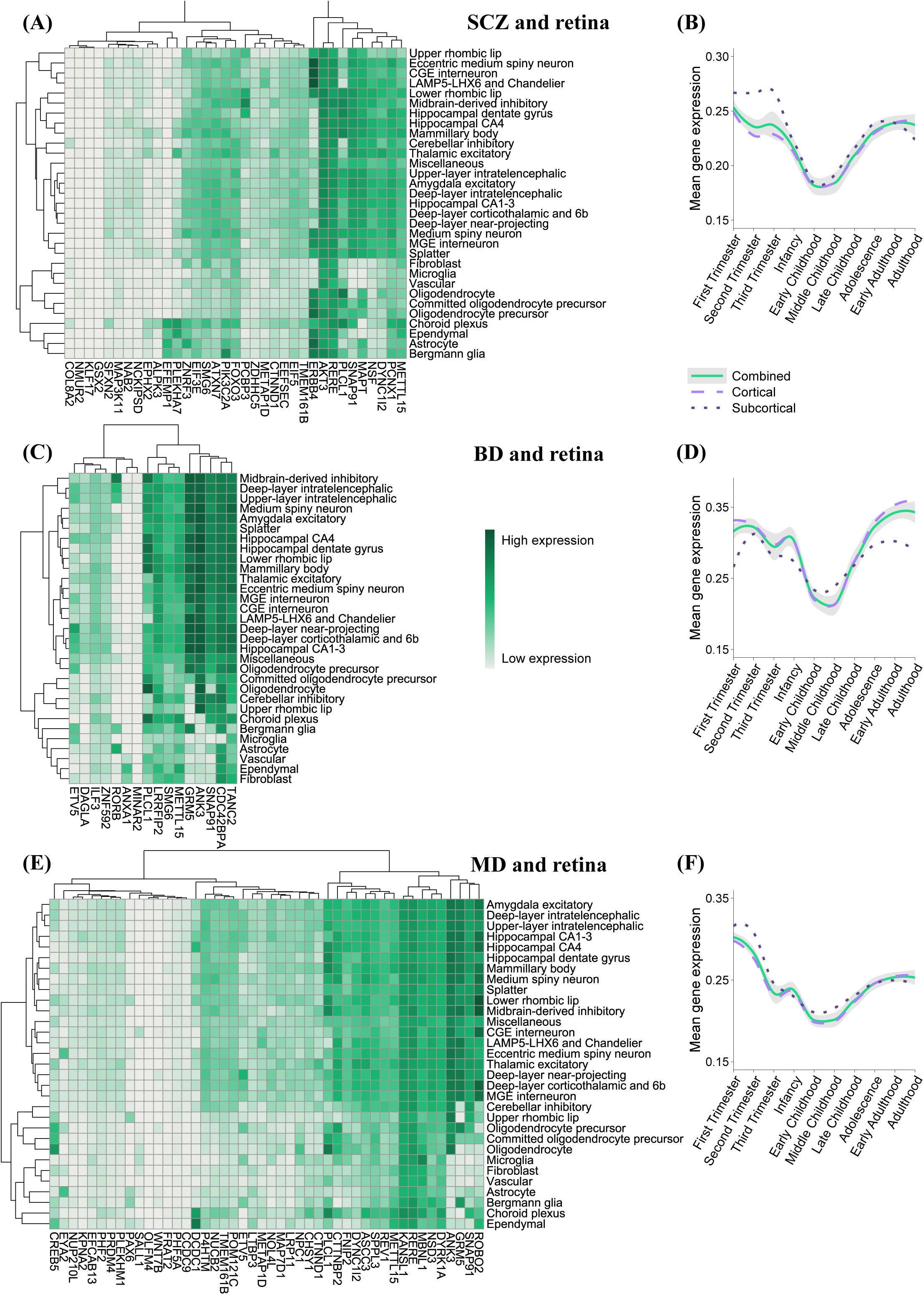
Analysis of genes associated with genetic variants shared between retinal traits and major psychiatric disorders. The expression analysis in the Whole Human Brain Atlas 10X RNA-seq dataset for genes shared between retinal traits and **(A)** SCZ, **(C)** BD, and **(E)** MD. Dendrograms and heatmaps illustrate the expression of these genes within clusters of neuronal and non-neuronal cells. Gene expression is indicated from low (light green) to high (dark green). Mean expression across lifespan periods for genes mapped to loci shared between retinal traits and **(B)** SCZ, **(D)** BD, **(E)** MD in cortical structures (purple line), subcortical structures (green line), and all structures combined (black line, with gray shading indicating the 95% confidence interval). Lines were fitted using nonlinear LOESS modeling.

The analysis of spatiotemporal expression indicated that genes shared between SCZ and retinal traits are predominantly expressed during the prenatal period and early adulthood (Figure 3B, Supplementary Figure 7). During the prenatal period, higher expression was observed in the striatum, amygdala, and hippocampus. In contrast, during early adulthood, higher expression was noted in all examined brain structures, with slightly increased expression in the primary visual cortex. Furthermore, the brain structure that consistently exhibited high expression of the studied genes across all examined life stages was the cerebellum. Genes mapped to the loci shared between retinal traits and BD are predominantly expressed in adulthood, with peak expression occurring in the primary visual cortex during early adulthood (Figure 3D). Genes mapped to the loci shared between retinal traits and MD are primarily expressed during the first two trimesters of pregnancy and early adulthood, and to a lesser extent in later adulthood (Figure 3F). This higher expression was consistent across all brain regions analyzed (Supplementary Figure 7).

Moreover, using PCNet v2.2 we evaluated interactions between protein products of the gene mapped to loci shared between retinal traits and MDSs (Figure 4 D-G). We observed a significantly higher number of protein-protein interactions for all three MPDs (*p*_nominal_=0.018, 0.023, 0.028 for SCZ, BD, MD, respectively, and *p*_adjusted_=0.028 for all three MPDs), when compared with randomly drawn protein-coding gene sets of the same-size. The numbers of genes interacting with each other within all three gene sets were not significantly higher than those observed in random protein-coding gene sets (all *p*_nomial_>0.05).

**Figure 4.**
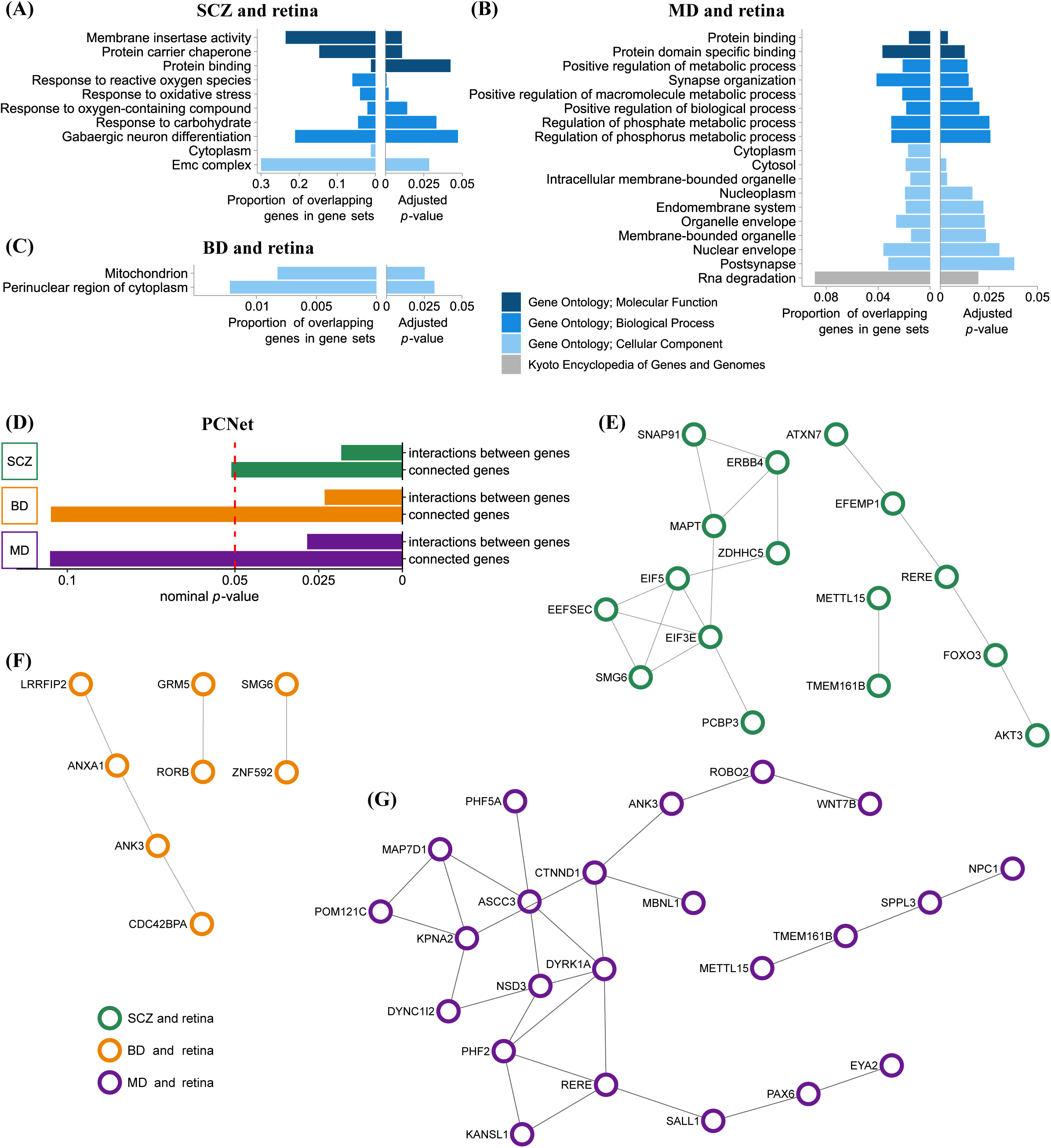
Analysis of genes associated with genetic variants shared between retinal traits and major psychiatric disorders. Gene set enrichment analyses for genes mapped to the loci shared between **(A)** retinal traits and schizophrenia (SCZ), **(B)** retinal traits and bipolar disorder (BD), and **(C)** retinal traits and major depression (MD). The color represents the gene-set category. All displayed results survived correction for multiple comparisons (adjusted *p*-value). **(D-G)** Interactome analysis using PCNet for genes mapped to genetic variants shared between retinal traits and major psychiatric disorders. **(D)** Enrichment analysis of interactions: number of interactions between mapped genes (interactions between genes), number of genes that interact with other mapped genes (connected genes). The vertical red dashed line represents the threshold for nominal significance. Interaction networks for the genes shared between retinal traits and: **(E)** schizophrenia (SCZ and retina), **(F)** bipolar disorder (BD and retina), **(G)** major depression (MD and retina).

Genes shared between retinal traits and SCZ exhibited enrichment in 10 gene ontology groups, those shared with BD were enriched in 2 gene ontology categories, whereas the genes shared with MD demonstrated enrichment in 17 gene ontology groups and the ‘RNA degradation’ gene set from KEGG (Figure 5 A-C).

**Figure 5.**
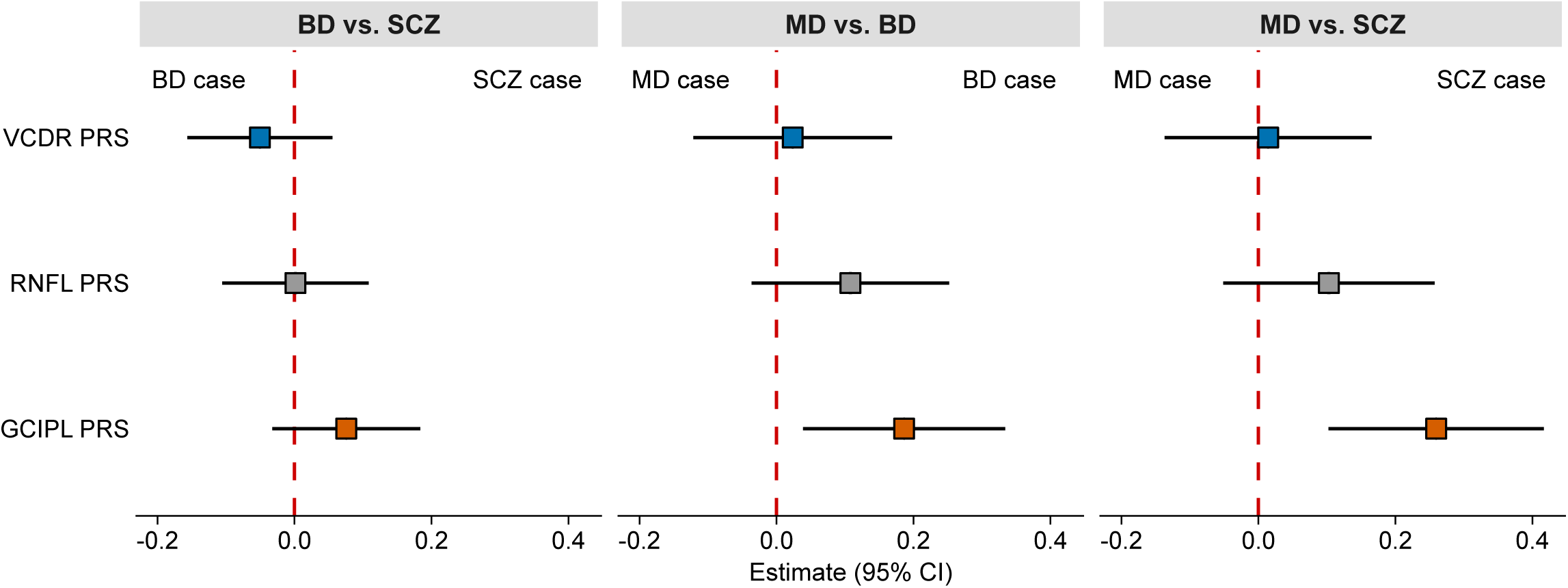
Association of PRS for retinal traits (ganglion cell inner plexiform layer thickness [GCIPL], retinal nerve fibre layer thickness [RNFL], vertical cup-disc ratio [VCDR]) with major depression (MD), bipolar disorder (BD) and schizophrenia (SCZ) compared to each of the other diagnostic categories derived from the logistic regression models. Error bars indicate 95% confidence intervals (95% CI) of the estimates. The vertical dashed red lines indicate an effect estimate equal to zero.

### Polygenic risk score analysis

We evaluated the association between retinal PRSs and MPD diagnoses in case-case logistic regression models (Figure 5). GCIPL PRS showed differential association with SCZ compared to MD cases (β=0.26; 95% CI, 0.10-0.42; *p*_nominal_=0.00121; *p*_adjusted_=0.0109). Moreover, we observed a nominally significant differential association of GCIPL PRS with BD cases compared to MD cases (β=0.19; 95% CI, 0.04-0.33; *p*_nominal_=0.0134), although this association did not remain significant after correction for multiple testing (*p*_adjusted_=0.060).

## Discussion

In our cross-trait genetic analysis, we estimated the genetic architecture shared between MPDs and retinal traits beyond genome-wide genetic correlations. Our results indicate that the retinal changes found in MPDs reflect a complex, disorder- and retinal trait-specific shared genetic architecture between retinal phenotypes and MPDs. The specificity of these overlapping genetic architectures translates into distinct developmental patterns of risk variant expression (e.g. supporting the two-stage developmental hypothesis of SCZ), as well as the involvement of divergent biological pathways across specific MPDs, with a role of GABAergic neurons in SCZ, mitochondrial systems in BD, and synaptic transmission in MD. Furthermore, as we have preliminarily demonstrated, the specificity of this shared genetic architecture may help differentiate between MPDs.

The MiXeR analysis showed that retinal traits, when compared to MPDs, are approximately 10-20 times less polygenic. This finding is consistent with previous observations indicating that MPDs, as complex phenotypes, are among the most polygenic human traits(28, 29). In contrast, less complex traits that are easier to define and measure (e.g., white blood cell counts, C-reactive protein level) are influenced by significantly fewer genetic variants(58, 59). We observed a gradient of differences in the proportion of shared variants with MPDs for individual retinal traits. The highest proportion of shared genetic variants with MPDs was detected in GCIPL, which shared an average of 65% of its trait-influencing variants. Conversely, the lowest proportion of shared variants was observed for VCDR, with an average of 20%. This may indicate that individual processes related to retinal structure are, to varying degrees, related to the genetic basis of MPDs. Further understanding of these processes may help identify clinically useful retinal biomarkers of MPDs.

Our conjFDR analysis indicates that the genetic variants shared between retinal traits and MPDs comprise a mixture of variants with both concordant and discordant effect directions, analogous to those observed in other human traits and disorders(28, 29, 60). This finding is also reflected in the low to moderate strength of genetic correlations of the shared genetic variants observed in the bivariate MiXeR models. The high degree of polygenicity of MPDs, which may reflect significant heterogeneity at both neurobiological and clinical levels(28), combined with a substantial genetic overlap between retinal traits and MPDs with mixed effect directions, reveals a complex relationship between the genetic underpinnings of retinal traits and MPDs. Due to this complexity, the phenotypic associations between a specific retinal trait and a major psychiatric disorder may vary among different neurobiologically distinct subgroups of patients. On one hand, this variability may contribute to the discrepancies observed in studies evaluating the link between retinal phenotypes and MPDs. On the other hand, this variability, along with the impact of administered treatments, the natural progression of the disease, and the presence of other comorbidities, further complicates efforts to elucidate the relationship between retinal traits and MPDs.

The gene enrichment analysis of genes mapped to variants shared between retinal traits and MPDs indicates that the biological underpinning of the relationship with retinal traits varies among different MPDs. In SCZ, the revealed involvement of biological processes, such as *the response to reactive oxygen species* and *oxidative stress*, as well as the role of the *protein carrier* chaperone, and the *endoplasmic reticulum membrane protein complex*, indicates the significance of oxidative stress and endoplasmic reticulum stress, which have previously been linked to the pathogenesis of SCZ(60–62) and retinal biology(63). Moreover, the observed involvement of *GABAergic neuron differentiation processes* confirms earlier findings regarding the role of GABAergic pathway in schizophrenia(64). Additionally, this involvement may suggest a role for amacrine cells, as the main retinal cell type releasing GABA(65, 66), in the relationship between schizophrenia and retinal traits. Boudriot et al. employed a different genetic analytical approach, identifying robust genetic associations between the risk of SCZ and amacrine cells(11). Enrichment analysis of the genes shared between BD and retinal traits has identified the involvement of the *mitochondrion*, which aligns with the mitochondrial dysfunction hypothesis of BD(67). Furthermore, this finding is consistent with observations regarding the crucial role of mitochondria in the proper functioning of the retina(68). Among the gene sets corresponding to genes shared between MD and retinal traits, we observed those related to synaptic function and RNA degradation. Functioning of synapses is regarded as a central element in the pathogenesis of MD(69, 70). Furthermore, retinal neurotransmission has been proposed as a marker for therapeutic response to antidepressant drugs that primarily target synaptic transmission(71). It has been demonstrated that disruptions in RNA degradation impact neuronal homeostasis, which may underlie neurodegenerative processes(72). Furthermore, our finding of increased connectivity for the mapped genes indicates that genes shaping retinal morphology and associated with MPD form functional networks. Understanding how these networks operate at a systems level has the potential to elucidate the mechanisms underlying the relationship between the retina and MPDs.

The results of expression analysis of genes mapped to loci shared between MPDs and retinal traits, across cell type clusters(47), indicate enrichment within neuronal cell clusters. This finding is consistent with previous studies that implicate neuronal gene expression in the pathogenesis of SCZ, BD, and MD(73, 74). Interestingly, our analysis of mapped genes expression in various brain structures across different life stages indicates differences between MPDs. For genes shared between retinal traits and SCZ, we find elevated expression levels in the amygdala, hippocampus, and striatum during the prenatal period. During early adulthood, consistent with the results of expression analyses in various neuronal cell clusters, we observe a similar gene expression pattern across all examined brain structures, with significantly higher expression noted in the primary visual cortex. These results align with the two-stage development hypothesis of SCZ(75) and observations suggesting a role for the amygdala, hippocampus(74), striatum(76), and primary visual cortex(77) in the disorder. Our results are in line with an association between retinal amacrine cells and SCZ during early developmental stages(11). Furthermore, they underscore Sullivan et al.’s observation regarding the need to analyse the influence of genetic variants on the development of SCZ over time, across different life stages, specific brain structures, and various cell types(73). Interestingly, the higher expression of genes identified as shared between retinal traits and BD only during (early) adulthood (unlike the genes shared between retinal traits and SCZ) appears to correlate with the less pronounced early developmental abnormalities in BD compared to SCZ(78). Additionally, this observation supports findings related to the occurrence of developmental abnormalities in early adulthood in BD. High expression of genes shared between retinal traits and MD during the prenatal period and early adulthood may reflect the crucial importance of these developmental stages in the emergence of susceptibility to MD, as well as time-related resilience to adverse life events(79, 80). Notably, comparing gene expression shared between retinal traits and MD with that of genes shared between retinal traits and SCZ may illuminate the specificity of susceptibility to early life adverse events in the contexts of SCZ and MD. In the case of MD, we observed expression limited to the first and second trimesters of pregnancy, distributed across all observed brain structures. In contrast, for SCZ, expression was evident throughout the prenatal period but was confined to select brain structures, as discussed above.

It is worth emphasizing that, although most of the genes mapped to variants associated with retinal traits were specific to MPDs, some of the mapped genes (e.g., *SNAP91*, *PLCL1*, *METAP1D*, *CTNND1*) were common to at least two MPDs. *SNAP91*, shared between GCIPL and the three MPDs, encodes synaptosome associated protein 91 that impacts synapsis functioning. It is predominantly expressed in the central nervous system and has been identified in the original GWAS for GCIPL, and it has also been linked to the pathogenesis of SCZ(81) and BD with mood-incongruent psychotic features(82). *PLCL1*, shared between VCDR and all three MPDs, encodes (inactive) phospholipase C-like 1, which is involved in the activity of the GABA receptor and has been linked to the neurodevelopmental mechanisms of SCZ(83). *METAP1D*, shared between SCZ, MD and RNFL, encodes mitochondrial methionyl aminopeptidase type 1D and has been linked with Leber’s hereditary optic neuropathy(84). *CTNND1*, shared among SCZ, MD, and RNFL, encodes catenin delta 1, responsible for neuronal migration and synaptogenesis. Previous studies have shown its association with the development of MD(85), vitreoretinopathy(86), and its influence on VCDR(7). The genes discussed above, whose genetic variants exhibit the same pattern of effects on all MPDs and a given retinal trait (e.g., the variants linked with *SNAP91* increase the risk of all these diseases: SCZ, BD, and MD, while reducing the thickening of the GCIPL), may be responsible for general susceptibility to MPDs. Whereas genetic variants with discordant effect directions on the risk of different MPDs may influence the risk for specific MPDs, such variants have been mapped to *RERE* and *ANK3*. *RERE* encodes a nuclear receptor co-regulator that regulates retinoic acid signaling(87). *RERE*-related neurodevelopmental disorders include brain and eye abnormalities(87). Furthermore, *RERE* has been linked to the pathogenesis of SCZ and MD(88, 89). *ANK3* encoding ankyrin 3 is mainly expressed in the nodes of Ranvier and the axon initial segment, where it stabilizes sodium channels by cross-linking them to spectrin-actin cytoskeleton proteins(90). It has been linked with neuropsychiatric disorders, including SCZ, and BD(90, 91).

Our PRS case-case comparison showed that the genetics of retinal traits may help to distinguish between different MPDs. These results are consistent with clinical observations indicating that specific changes in retinal traits structure and function may be useful in differentiating between MPDs(8, 21). Furthermore, together with other analyses from our study, they indicate that it is genetic risk variants that may underlie the differences in retinal structure and functioning observed in the clinic.

The results of our study allow us to suggest future research on the relationship between retinal traits and MPDs. Firstly, due to the complexity of the genetic architecture shared between retinal traits and MPDs, future studies should adequately address the issue of variability (heterogeneity) within a given disorder by carefully selecting and describing the genetic and clinical characteristics of the studied groups. It can be assumed that within one diagnostic entity, specific patient groups may differ in their retinal phenotype characteristics. Importantly, histological studies have reached similar conclusions: specific morphological alterations in brain structure (including components of the visual system) -correlate with the symptom profiles of subtypes of a given MPD(1). Secondly, the time-varying gene expression profiles shared between retinal traits and MPDs underscore the necessity of incorporating the time factor (developmental phase) when assessing the influence of environmental risk factors, such as infections, on the relationship between psychiatric disorders and retinal phenotypes. Thirdly, our study provides further evidence indicating that, due to the complexity and specificity of the relationships between retinal and psychiatric traits, the continued development of retinal phenotype assessment techniques, combined with tools for analyzing large-scale and complex data sets, such as artificial intelligence, can yield essential diagnostic and prognostic biomarkers for psychiatry. Fourthly, although these remarks should be considered preliminary, since the genetic variants shared between MPDs and retinal traits constitute only a fraction of the genetic basis of MPDs, considering the genetic perspective, we may expect that retinal traits could serve as tentative biomarkers for individual processes related to the pathogenesis of MPDs or help differentiate specific patient subgroups, rather than assessing the overall risk of MPDs.

The are some limitations of our study. In our analysis, we did not examine both eyes separately for sidedness and dominance(13). Considering this perspective, future studies may provide additional support for our conclusions regarding the complex relationship between retinal traits and MPDs. Furthermore, the inclusion of non-European populations in future studies is necessary to validate the generalizability of our results.

Our findings of overlapping genetic architecture across MPDs and retinal traits revealed novel insights into the molecular underpinnings of clinically observed relationships between these phenotypes. Furthermore, they highlighted disorder-specific mechanisms and their developmental trajectories. Finally, our results indicate the potential of leveraging retinal traits as clinical biomarkers in psychiatry.

## Data Availability

All data produced in the present work are contained in the manuscript.

## Acknowldgements

The authors were funded by the Research Council of Norway (grants #324252, #326813, #334920, #324499, #223273, #248778), the European Union’s Horizon 2020 Research and Innovation Programme (#847776 and #964874), the European Union’s Horizon 2020 research and innovation programme under the Marie Sklodowska-Curie grant (#801133), the European Union grant (#101057429, environMENTAL), the National Institutes of Health (#U24DA041123, #R01AG076838, #OT2HL161847 and #U24DA055330), the EEA and Norway grant (#EEA-RO-NO-2018-0573), KG Jebsen Stiftelsen, and the South-East Norway Regional Health Authority (#2022-087, #2022073), Wellcome Leap, CARE Program (“FEMA-AD”). This work also used the TSD (Tjeneste for Sensitive Data) facilities, owned by the University of Oslo, operated and developed by the TSD service group at the University of Oslo, IT-Department (USIT), using resources provided by UNINETT Sigma2 – the National Infrastructure for High Performance Computing and Data Storage in Norway. The funders had no role in study design, data collection and analysis, decision to publish or preparation of the manuscript.

