## Supplementary Figures for "Molecular Underpinnings of Retinal Traits Shared with Major Psychiatric Disorders": retina_supplement_09_2026.docx

**Table of contents**

Supplementary Figure 1 ………………………………………………………………….…….3

Supplementary Figure 2 ………………………………………………………………….…….4

Supplementary Figure 3 ………………………………………………………………….…….5

Supplementary Figure 4 ………………………………………………………………….…….6

Supplementary Figure 5 …………………………………………………………………….….7

Supplementary Figure 6 ………………………………………………………………………..8

Supplementary Figure 7 …………………………………………………………………..…....9

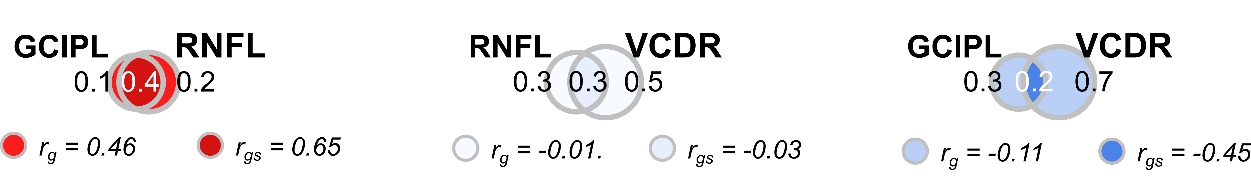

**Supplementary Figure 1.** Venn diagrams illustrating MiXeR-modeled genetic overlap, genome-wide genetic correlation, and genetic correlation of shared variants between retinal traits. The size of the circles reflects the polygenicity of each trait. The unique and shared regions of the Venn diagrams show the numbers of unique and shared trait-influencing variants, respectively, with the number of trait-influencing variants shown in thousands. Genome-wide genetic correlations (rg) are depicted below each Venn diagram and are represented by coloring the unique parts of each diagram. Genetic correlations of shared variants (rgs) are depicted below each Venn diagram and are represented by coloring the shared part of each diagram. The colours illustrate correlations, ranging from 1 (dark red) to -1 (dark blue). Additionally, the SNP-based heritability (h²_SNP_) and total polygenicity (polygenicity90) are presented for each trait. Ganglion cell inner plexiform layer thickness (GCIPL); retinal traits such as retinal nerve fibre layer thickness (RNFL); and vertical cup-disc ratio (VCDR).

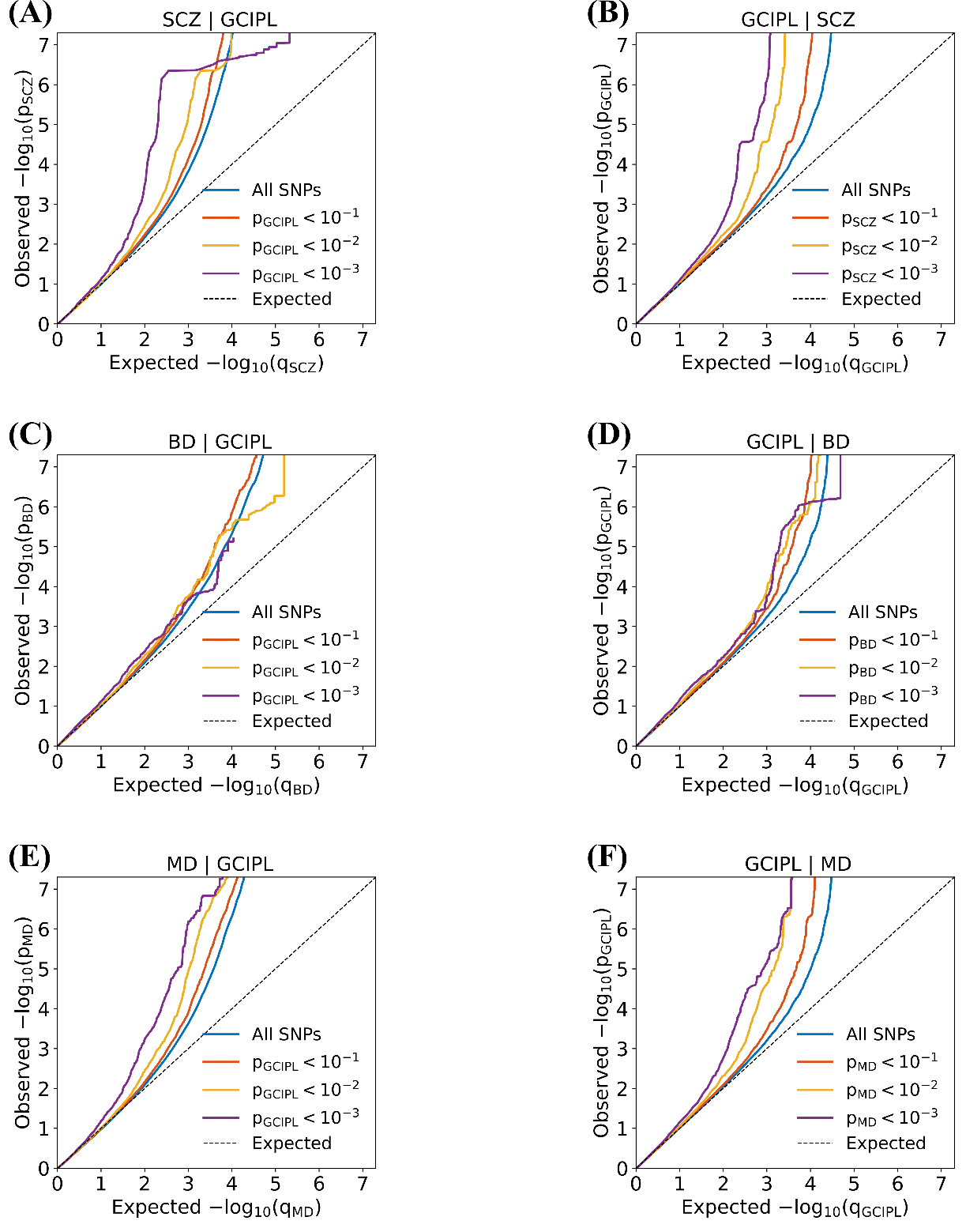

**Supplementary Figure 2.** Conditional QQ plots show cross-trait enrichment of ganglion cell inner plexiform layer thickness (GCIPL) and major psychiatric disorders (MPDs) at different threshold values. A) Schizophrenia (SCZ) conditioned on GCIPL (SCZ | GCIPL ), B) GCIPL conditioned on SCZ (GCIPL | SCZ), C) bipolar disorder (BD) conditioned on GCIPL (BD | GCIPL), D) GCIPL conditioned on BD (GCIPL | BD), E) major depression (MD) conditioned on GCIPL (MD | GCIPL), F) GCIPL conditioned on MD (GCIPL | MD).

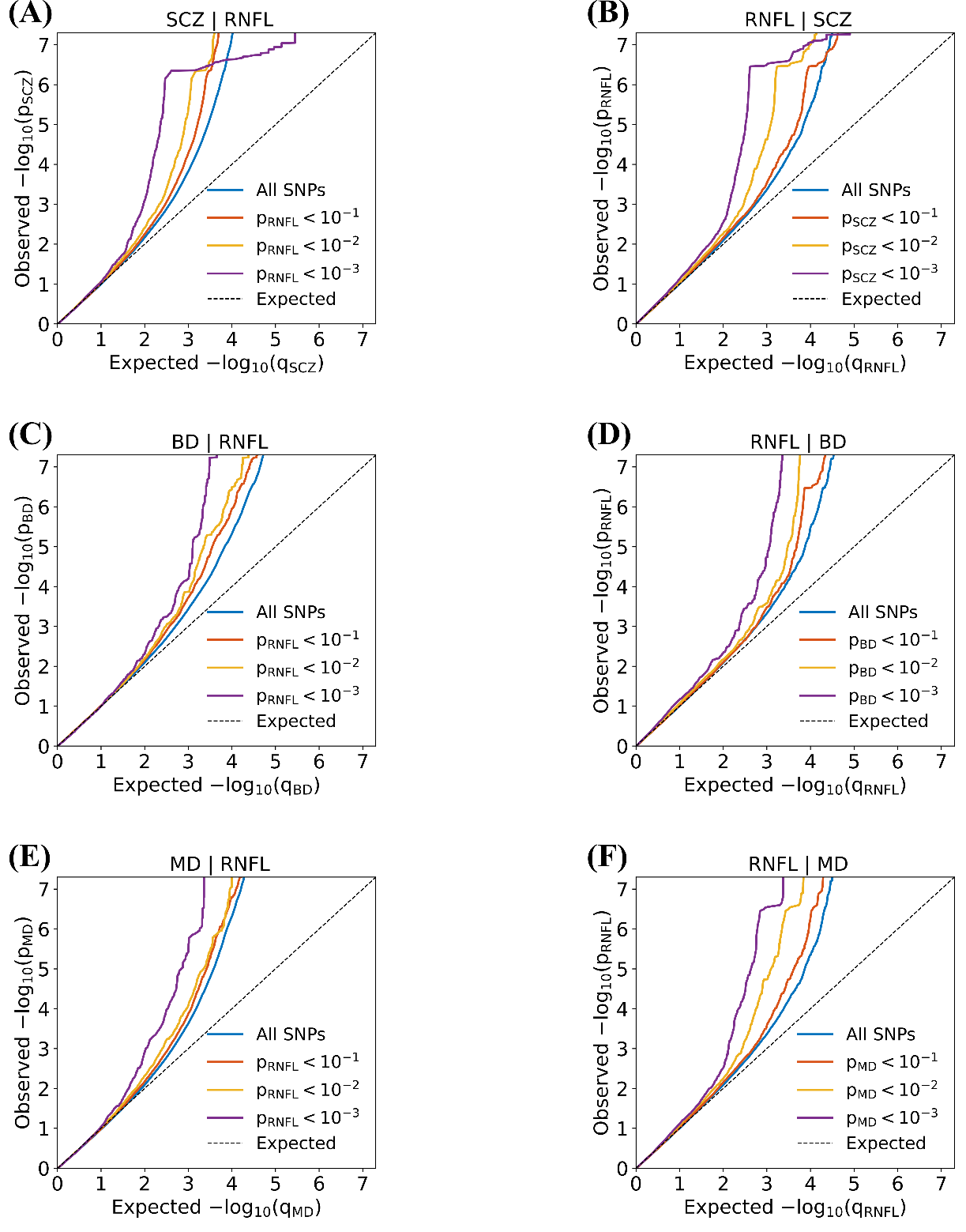

**Supplementary Figure 3.** Conditional QQ plots show cross-trait enrichment of retinal nerve fiber layer thickness (RNFL) and major psychiatric disorders (MPDs) at different threshold values. A) Schizophrenia (SCZ) conditioned on RNFL (SCZ | RNFL ), B) RNFL conditioned on SCZ (RNFL | SCZ), C) bipolar disorder (BD) conditioned on RNFL (BD | RNFL), D) RNFL conditioned on BD (RNFL | BD), E) major depression (MD) conditioned on RNFL (MD | RNFL), F) RNFL conditioned on MD (RNFL | MD).

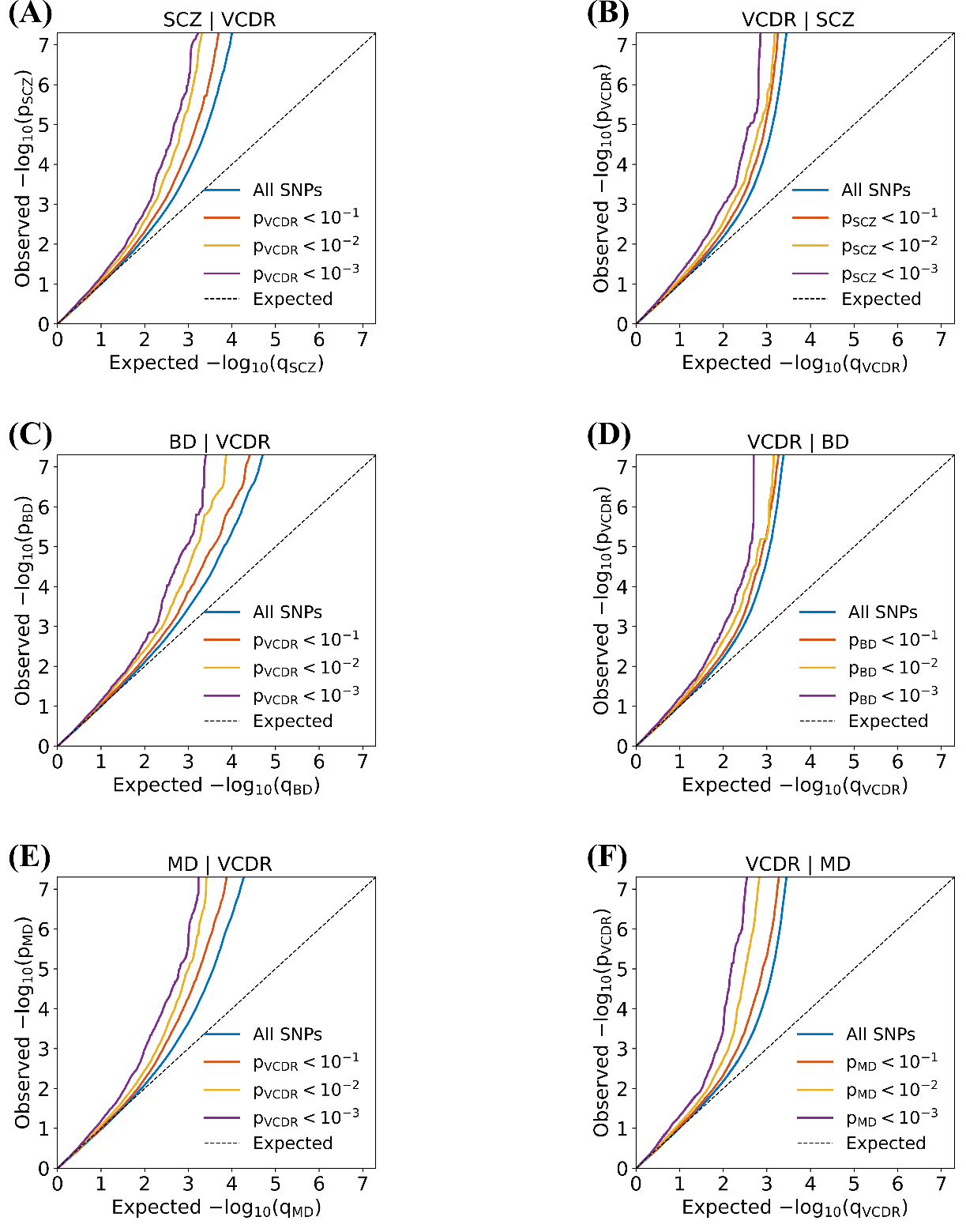

**Supplementary Figure 4.** Conditional QQ plots show cross-trait enrichment of vertical cup-disc ratio (VCDR) and major psychiatric disorders (MPDs) at different threshold values. A) Schizophrenia (SCZ) conditioned on VCDR (SCZ | VCDR ), B) VCDR conditioned on SCZ (VCDR | SCZ), C) bipolar disorder (BD) conditioned on VCDR (BD | VCDR), D) VCDR conditioned on BD (VCDR | BD), E) major depression (MD) conditioned on VCDR (MD | VCDR), F) VCDR conditioned on MD (VCDR | MD).

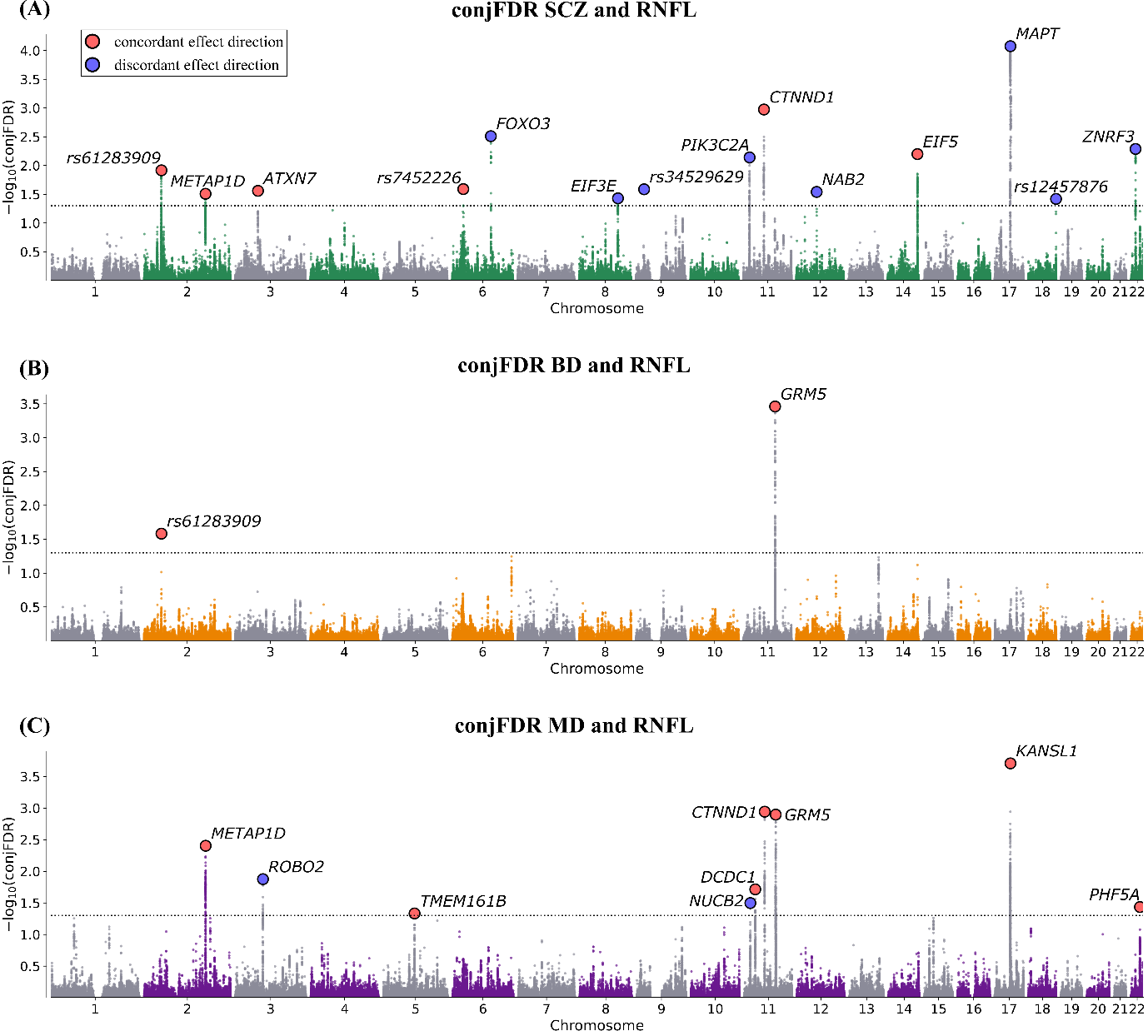

**Supplementary Figure 5.** The conjFDR Manhattan plots illustrate common genetic variants that are jointly associated with (A) schizophrenia (SCZ) and retinal nerve fibre layer thickness (RNFL), (B) bipolar disorder (BD) and RNFL, and (C) major depression (MD) and RNFL at conjunctional false discovery rate (conjFDR) < 0.05. The y-axis shows the −log10 transformed conjFDR. Chromosomal position is presented along the x-axis. The threshold for significant shared associations (conjFDR < 0.05) is represented by the horizontal dotted line. Lead SNPs are indicated by a black perimeter. Lead SNPs with concordant effect directions for both traits are depicted in red, whereas those with discordant effect directions are depicted in blue.

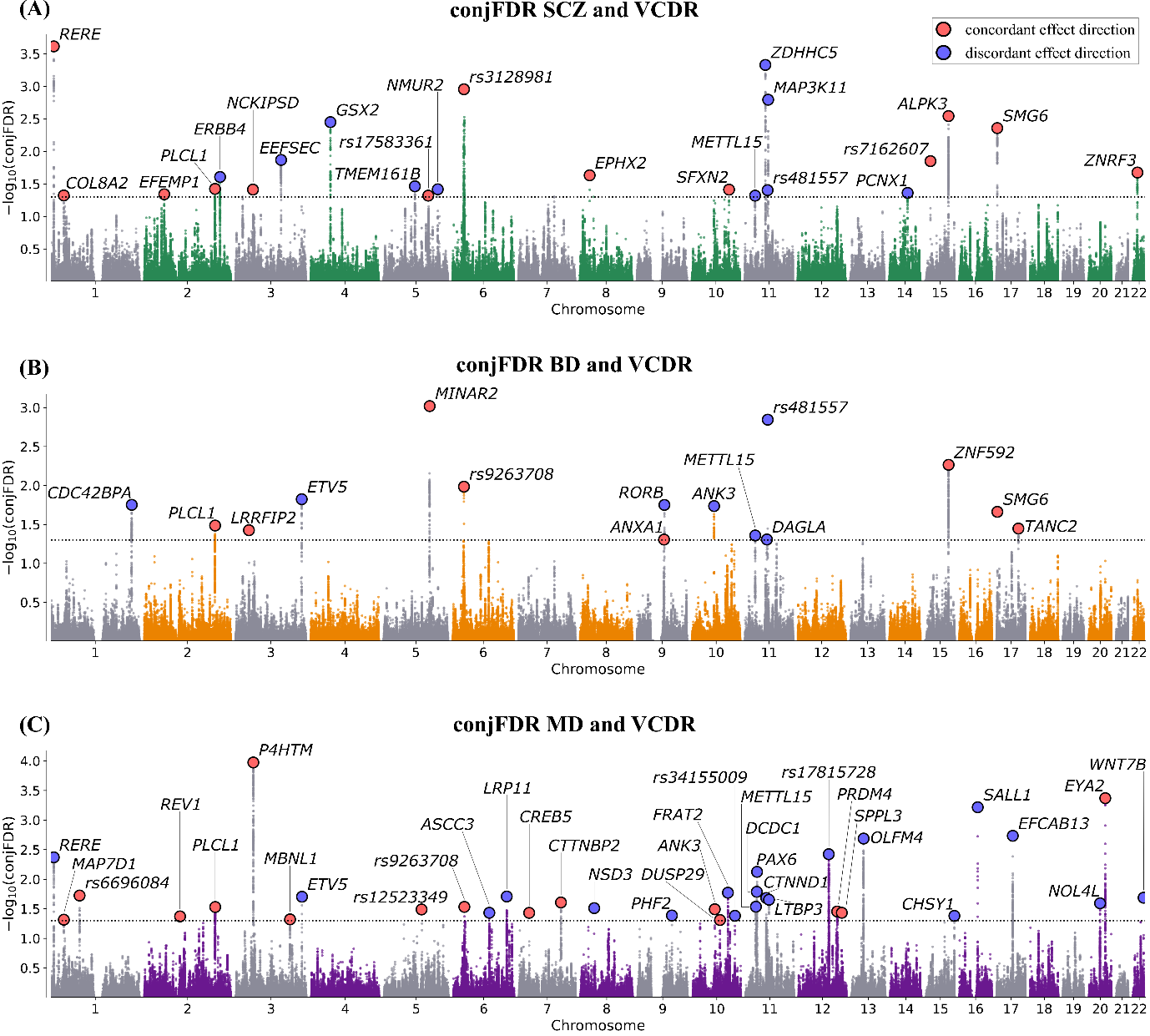

**Supplementary Figure 6.** The conjFDR Manhattan plots illustrate common genetic variants that are jointly associated with **(A)** schizophrenia (SCZ) and vertical cup-disc ratio (VCDR), **(B)** bipolar disorder (BD) and VCDR, and **(C)** major depression (MD) and VCDR at conjunctional false discovery rate (conjFDR) < 0.05. The y-axis shows the −log10 transformed conjFDR. Chromosomal position is presented along the x-axis. The threshold for significant shared associations (conjFDR < 0.05) is represented by the horizontal dotted line. Lead SNPs are indicated by a black perimeter. Lead SNPs with concordant effect directions for both traits are depicted in red, whereas those with discordant effect directions are depicted in blue.

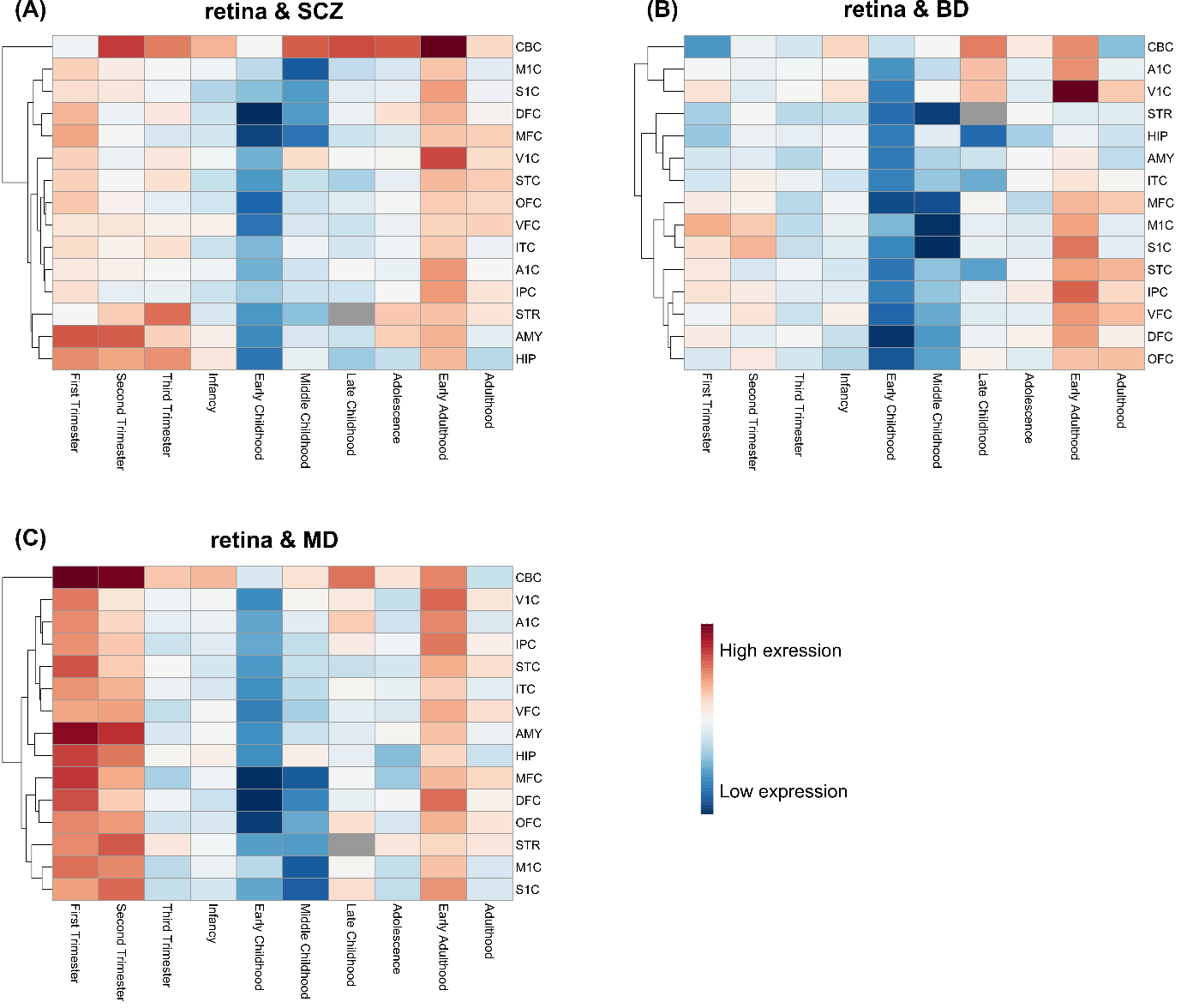

**Supplementary Figure 7. Spatiotemporal expression of genes associated with genetic variants shared between retinal traits and major psychiatric disorders.** A dendrogram and heatmap illustrating the spatiotemporal gene expression of genes mapped for (A) retinal traits and schizophrenia (SCZ), (B) retinal traits and bipolar disorder (BD), as well as (C) retinal traits and major depression (MD), using RNA sequencing data from BrainSpan across 10 developmental periods (columns) and 15 brain regions (rows).Gene expression is indicated from high (red) to low (blue); amygdala (AMY), cerebellum (CBC), dorsolateral prefrontal cortex (DFC), hippocampus (HIP), inferior parietal cortex (IPC), medial prefrontal cortex (MFC), superior temporal cortex (STC), striatum (STR), auditory cortex (A1C), primary sensory (S1C), primary motor (M1C), primary visual (V1C), inferior temporal (ITC), ventrolateral prefrontal (VFC), orbitofrontal cortices (OFC).
